# Mining transcriptomics and clinical data reveals ACE2 expression modulators and identifies cardiomyopathy as a risk factor for mortality in COVID-19 patients

**DOI:** 10.1101/2020.10.20.20216150

**Authors:** Navchetan Kaur, Boris Oskotsky, Atul J. Butte, Zicheng Hu

## Abstract

Angiotensin-converting enzyme 2 (*ACE2*) is the cell-entry receptor for SARS-CoV-2. It plays critical roles in both the transmission and the pathogenesis of the coronavirus disease 2019 (COVID-19). Comprehensive profiling of ACE2 expression patterns will help researchers to reveal risk factors of severe COVID-19 illness. While the expression of ACE2 in healthy human tissues has been well characterized, it is not known which diseases and drugs might modulate the ACE2 expression. In this study, we developed GENEVA (GENe Expression Variance Analysis), a semi-automated framework for exploring massive amounts of RNA-seq datasets. We applied GENEVA to 28,6650 publicly available RNA-seq samples to identify any previously studied experimental conditions that could directly or indirectly modulate ACE2 expression. We identified multiple drugs, genetic perturbations, and diseases that modulate the expression of ACE2, including cardiomyopathy, HNF1A overexpression, and drug treatments with RAD140 and Itraconazole. Our unbiased meta-analysis of seven datasets confirms ACE2 up-regulation in all cardiomyopathy categories. Using electronic health records data from 3936 COVID19 patients, we demonstrate that patients with pre-existing cardiomyopathy have an increased mortality risk than age-matched patients with other cardiovascular conditions. GENEVA is applicable to any genes of interest and is freely accessible at http://genevatool.org.

## Introduction

Coronavirus disease 2019 (COVID-19) is an infectious disease caused by severe acute respiratory syndrome coronavirus 2 (SARS-COV-2). The World Health Organization (WHO) declared the COVID□ 19 outbreak a pandemic on March 11, 2020. As of October 5, 2020, there have been 34.8 million recorded COVID-19 cases and over 1 million deaths^1^.

Angiotensin-converting enzyme 2 (ACE2) is the cell-entry receptor for SARS-CoV-2^2^. The binding between ACE2 and spike (S) protein of SARS-COV-2 initiates the viral entry into target cells. ACE2 plays key roles in both the transmission and pathogenesis of SARS-CoV-2, as demonstrated by the following lines of evidence: 1) SARS-CoV-2 fails to infect the lung-derived cell line A549 in the absence of ACE2 expression. The infection is restored after overexpressing ACE2 in the cell line^3^. 2) SARS-CoV-2 fails to infect wild type mice but can infect and cause pneumonia in transgenic mice expressing human ACE2^4,5^. 3) COVID19 related tissue damages are detected in organs with ACE2 expression, including lungs, intestines, colons, and hearts^6–8^. 4) ACE2 expression is increased in the lungs of patients with comorbidities associated with severe COVID-19, suggesting that the level of ACE2 expression is associated with disease severity^9^. Taking these lines of evidence together, it is crucial to comprehensively characterize the ACE2 expression in human tissues.

To comprehensively profile the expression patterns of ACE2, we not only need to characterize its expression in healthy tissues but also identify diseases, drugs and genetic perturbations that modulate ACE2 expression. The expression of ACE2 in healthy human tissues has been well characterized by resources such as the Human Cell Atlas and GTEx, with the highest expression detected in intestine, testis, lung, cornea, heart, kidney, and adipose tissues^10,11^. However, it is still not clear which diseases and drugs modulate the ACE2 expression. Since ACE2 expression is tightly associated with the pathogenicity of SARS-COV-2, identification of ACE2 modulating conditions will help us reveal and explain risk factors of severe illness from COVID-19.

RNA-sequencing data profiles the full transcriptome of samples. Currently, more than 200,000 human RNA-seq samples are publicly available, providing an unprecedented opportunity for us to examine ACE2 expression in different human cell types under a variety of conditions and treatments. Data harmonization efforts such as ARCHS4 have uniformly preprocessed the RNA-seq data, making them readily available for analysis^12^. However, fully automated analysis of these datasets faces two main obstacles. First, the metadata are non-standardized and are often unstructured, making it difficult to extract experimental conditions from the studies. Second, experimental designs are highly variable. While some studies adopt the simple control-versus-treatment design, other studies are more complicated, involving multiple time points, combination treatments, or stratified cohorts. The heterogeneous design makes it difficult to analyze the datasets using a single statistical model.

Multiple tools have been made to analyze transcriptomics data, including CREEDS^13^, scanGEO^14^, GEM-TREND^15^, StarGEO^16^, SIGNATURE^17^, SPIED^18^, Cell Montage^19^, ProfileChaser^20^, ExpressionBlast^21^ and SEEK^22^. However, the existing tools have several limitations, preventing them from fully exploring the publicly available RNA-sequencing resources. First, some of the tools annotate the metadata manually and are unable to cover the large number of datasets currently available. Second, the tools focus on differential expression analysis between two groups (e.g., control versus treatment), preventing them from analyzing studies with more complex study designs.

In this study, we developed GENEVA (GENe Expression Variance Analysis), a semi-automated framework for exploring public RNA-seq datasets. For a given gene, GENEVA identifies the most relevant datasets by analyzing the variance of the gene expression. GENEVA visualizes the relevant datasets for detailed manual analysis. GENEVA is scalable and is agnostic to study designs. Using GENEVA, we identified multiple drugs, genetic perturbations, and diseases that modulate the expression of ACE2, including cardiomyopathy, HNF1A over-expression,and drug treatments with RAD140 and Itraconazole. Our in-depth meta-analysis of seven datasets reveals increased ACE2 expression in all cardiomyopathy categories. By analyzing the clinical data of 3936 COVID19 patients at UCSF hospital, we demonstrate that patients with pre-existing cardiomyopathy have an increased mortality risk than other patients, including propensity score-matched patients with other cardiovascular conditions.

## Results

### Analysis of 286650 RNA-seq samples reveals complex transcriptional networks of ACE2

Our study leverages human RNA-sequencing data from the ARCHS4 project, containing 286,650 uniformly pre-processed data from 9,124 Gene Expression Omnibus (GEO) series^12^. The large number of RNA-sequencing samples provide an unprecedented resource for studying the expression of ACE2 in different human cell types under a variety of conditions and treatments.

We first characterized the transcriptional networks of ACE2 using all 286,650 samples. We calculated the Pearson correlation between ACE2 and all other human genes. Because of the large sample size, most of the correlations are statistically significant, even after multiple testing adjustments. Therefore, we focused on the correlation coefficients themselves as a measure of effect size, rather than the p values or significance. While most of the genes have correlation coefficients near 0, a small set of genes are highly correlated with ACE2 (Supplemental Fig. 1A). Among 35,238 human genes, 85 (0.24%) genes have correlations higher than 0.5. The top correlated genes include FABP2, MEP1B, and transcription factors such as HNF4G (Supplemental Fig. 1B and C). We examined their expression in different human tissues using GTEx data. We found that these genes are highly expressed in colons and small intestines, an expression pattern similar to ACE2. Similarly, we see high correlations between ACE2 and multiple pathways related to the digestive process (Supplemental Fig. 1D). These results indicate that the correlations are dominated by the overall transcriptomic difference between the digestive system and other human organs.

**Figure 1:**
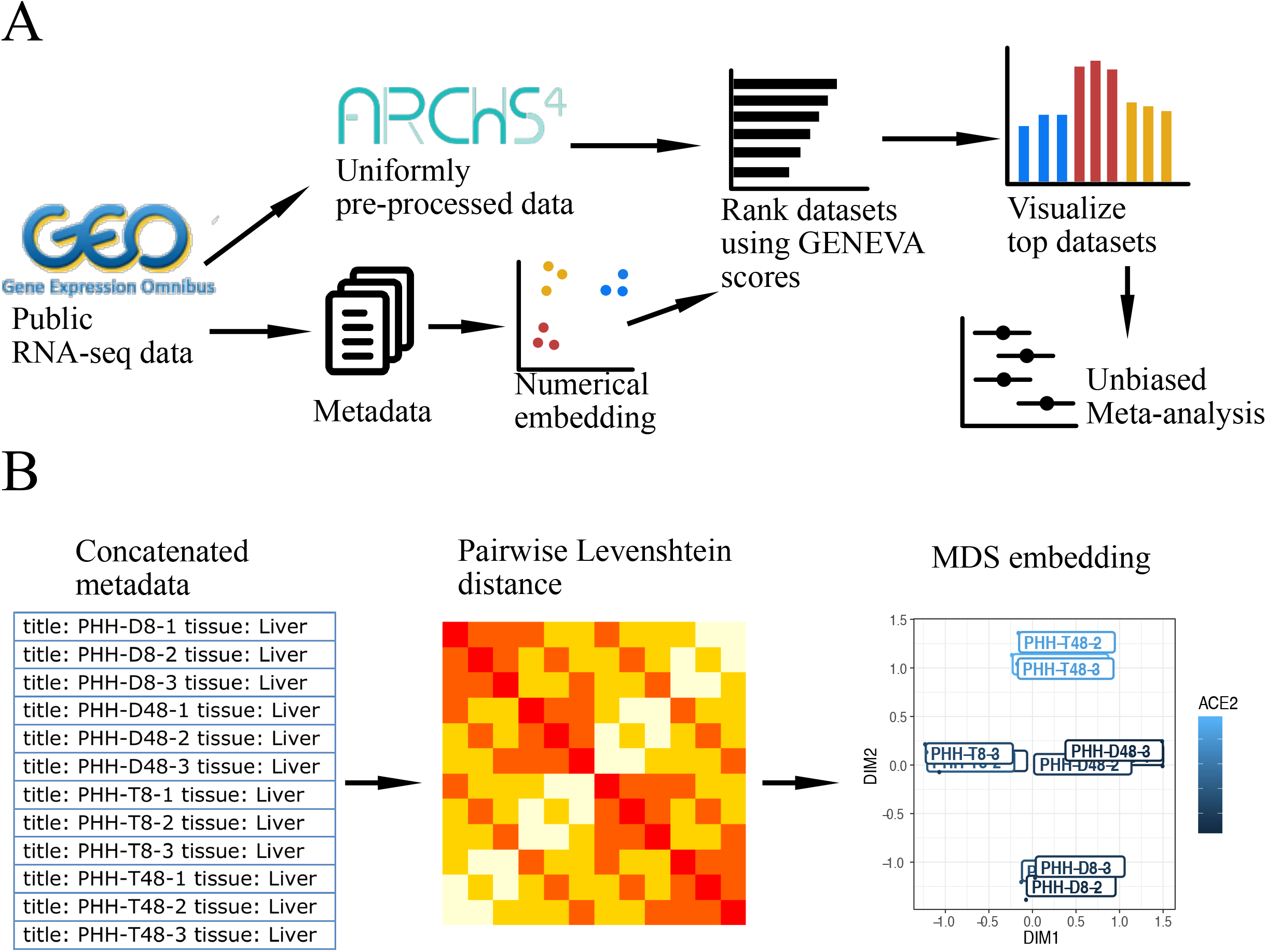
Mining RNA-seq data using Gene Expression Variance Analysis (GENEVA). (A) The workflow of GENEVA. (B) The procedure for embedding metadata into numerical vectors.

To avoid our analysis being dominated by known cellular or organ differences, we evaluated the correlations between ACE2 and other human genes within each RNA-seq dataset. We then calculated the mean and variances of the correlations across all datasets, weighted by the sample sizes. Our analysis reveals a positive relationship between the mean and variance of the correlation coefficients (Supplemental Fig. 1E). While some genes have a high average correlation with ACE2, their correlation with ACE2 is highly variable in individual datasets. The results suggest against a common transcriptional network around the ACE2 gene. Rather, ACE2 is co-regulated with different sets of genes under different conditions.

We adjusted for the variances by calculating the standardized correlation coefficient, defined as the mean correlation divided by the standard deviation of the correlation. The standardized correlation coefficient allows us to prioritize genes with relatively conserved correlations with ACE2 across all studies. The top genes include MYO7B, PDZK1 and transcription factors such as HNF1A (Supplemental Fig. 1 F and G). Pathway analysis identified many metabolic processes to be correlated with ACE2 expression. The findings are consistent with previous observations that ACE2 is involved in glucose metabolism and energy stress responses^23,24^.

Our analysis identified three transcription factors in the hepatocyte nuclear factor family, including HNF4G, HNF1A, and HNF4A (Supplemental Fig. 1 C and G). HNF4G has the highest overall correlation with ACE2 while HNF1A has the highest standardized correlation coefficient within studies. We then tested the causal relationship between the transcription factors and ACE2 expression. We identified two RNA-seq datasets that compared human cells with or without genetic perturbation of HNF4G and HNF1A. While HNF1G is positively correlated with ACE2 expression (Supplemental Fig. 1 C), overexpression of HNF1G does not lead to significantly increased ACE2 expression (Supplemental Fig. 1I). Rather, there is a trend of reduction in ACE2 expression. HNF1A overexpression leads to increased ACE2 expression and HNF1A knockdown reduced ACE2 expression (Supplemental Fig. 1J) in LNCaP cells, a prostate cancer cell line. The result is consistent with the positive correlation between HNF1A and ACE2 (Supplemental Fig. 1G). A previous study showed that HNF1A induces ACE2 in pancreatic islets^25^. Our result in a prostate cancer cell line further confirmed the role of HNF1A in regulating ACE2. However, it should be noted that HNF1A and ACE2 are not correlated in all RNA-seq datasets (Supplemental Fig. 1K), demonstrating the complexity of ACE2 regulation in different tissues.

### Gene expression variance analysis reveals diseases and therapeutics that modulate the expression of ACE2

Next, we hope to identify conditions that modulate the expression of ACE2. We developed a computational framework named GENEVA (Gene Expression Variance Analysis) to identify the most relevant datasets for visualization and detailed manual analysis (Fig. 1A and Methods). GENEVA prioritizes the datasets that have a large variance of ACE2 expression. The rationale is that datasets with large ACE2 variance are likely to contain conditions that modulate the ACE2 expression. At the same time, GENEVA controls for the overall heterogeneity of the samples to prioritize datasets in which ACE2 is specifically modulated by experimental conditions rather than due to tissue type differences. In addition, GENEVA embeds the meta-data into numerical space and prioritizes datasets with high correlations between ACE2 expression and the metadata (Fig. 1B). This allows GENEVA to identify datasets in which ACE2 is regulated by experimental conditions rather than randomness or unexplained factors. While our study focuses on ACE2 and its role in COVID-19 disease, GENEVA is applicable to all genes. We created a web application that allows researchers to apply GENEVA to their gene of interest [http://genevatool.org].

We tested the significance of the GENEVA scores using a permutation procedure. We randomly shuffle the samples across studies to generate a null distribution. We compared each GENEVA score to the null distribution to calculate the p-value. We adjusted for multiple-testing using the false discovery rate (FDR) method^26^. We identified 27 significant datasets with FDRs less than 0.05 (Table 1). Interestingly, GENEVA identified HNF1A as an ACE2 modulator, which was also identified in our correlation analysis (Supplemental Fig. 1J). GENEVA additionally identified multiple drugs and diseases that modulate the ACE2 expression, revealing potential risk factors for severe illness from COVID-19.

**Table 1:**
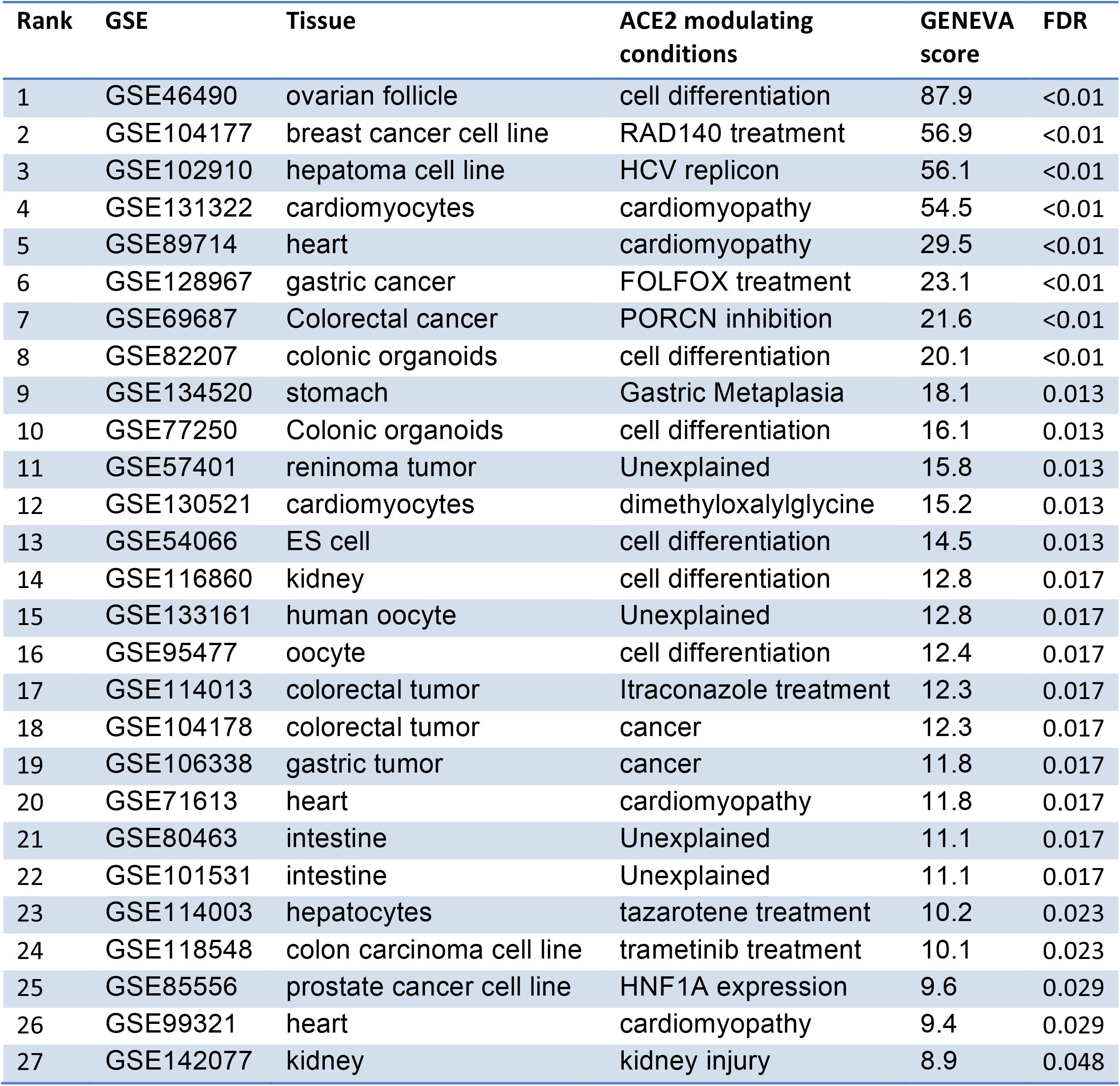
RNA-seq datasets with GENEVA scores of ACE2 expression that are statistically significant.

Here, we highlight three ACE2 modulating conditions, manually picked based on their effect on ACE2 expression and their potential impact on public health. Data from GSE89714 show up-regulated expression of ACE2 in hypertrophic cardiomyopathy (Fig. 2 A, B). Hypertrophic cardiomyopathy is the most common inherited heart disease, affecting an estimated 15,188,000 individuals (0.2%) worldwide^27^. Our finding is consistent with an increased death rate in COVID-19 patients with heart conditions^28–30^ and suggests that higher ACE2 expression can contribute to the increased risk. Data from GSE104177 showed that RAD140, a selective androgen receptor modulator, induces ACE2 expression in human breast cancer xenografts (Fig. 2 C, D). Data from GSE114013 show that Itraconazole, an antifungal drug, up-regulates ACE2 expression in two colorectal cancer cell lines, HT55 and SW948 (Fig. 2 E, F). These findings suggest that these drugs should be studied with respect to ACE2 expression in lung and cardiac cells and tissues, and that patients on these drugs should be studied closely during the pandemic. If these subsequent studies do continue to suggest this effect on increasing ACE2 expression, heightened caution could be warranted when using these drugs during the COVID-19 pandemic.

**Figure 2:**
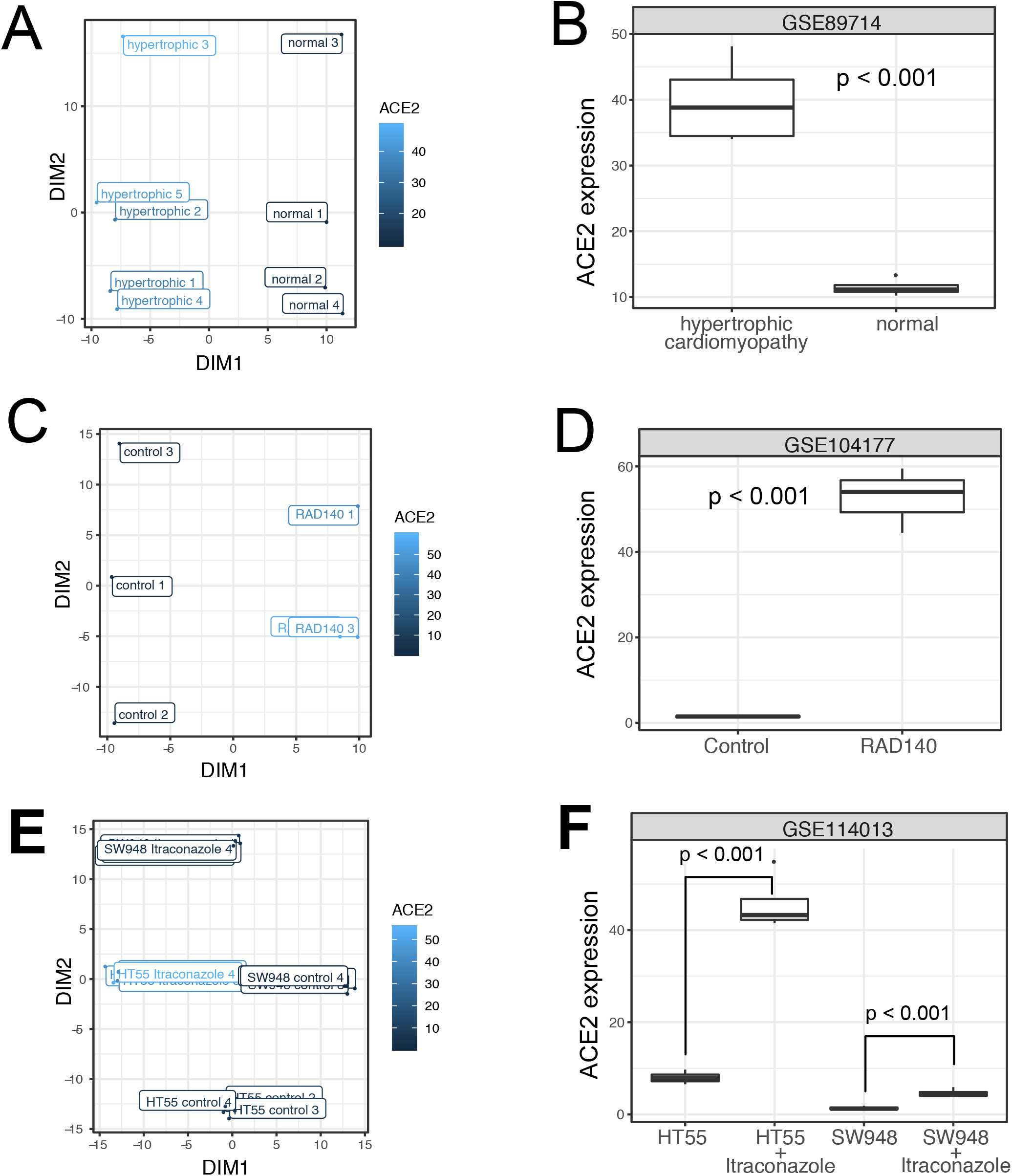
Highlighted conditions that modulate ACE2 expression. (A-B) Plots showing data from a study with GEO accession GSE89714. (A) A scatter plot showing the association between ACE2 expression and the embedded metadata. All metadata fields are concatenated for embedding. The plot only shows the sample title as labels. The labels are jittered to avoid perfect overlap. The color code represents the ACE2 expression level. Box plot showing the ACE2 expression in normal hearts and hearts with hypertrophic cardiomyopathy. (C-D) Plots visualizing data from a study with GEO accession GSE104177, showing the ACE2 expression in breast cancer xenografts with or without RAD140 treatment. (E-F) Plots visualizing data from a study with GEO accession GSE114013, showing the ACE2 expression in two prostate cancer cell lines with or without Itraconazole treatment.

### Unbiased meta-analysis shows ACE2 up-regulation in all types of cardiomyopathy

GENEVA prioritizes datasets with large variances in ACE2 expression. However, the procedure may introduce bias, as studies with small ACE2 variations are ignored. Consider an example in which multiple studies have profiled the effect of a drug. Some studies show that the drug up-regulates ACE2 while other studies show that the drug has no effect on ACE2. The effect of the drug will be overestimated if a researcher only includes the studies with positive results. Therefore, after the GENEVA analysis, unbiased meta-analyses are required to confirm the findings.

We performed a comprehensive search for datasets related to the three highlighted conditions, including cardiomyopathy, itraconazole treatment, and RAD140 treatment. We did not find additional datasets related to itraconazole and RAD140 treatment. For cardiomyopathy, we identified a total of 7 datasets. We performed a meta-analysis using a mixed-effect model, taking data from all 7 datasets into account. The result confirmed that ACE2 expression is significantly elevated in heart tissue samples from cardiomyopathy patients (p-value < 0.001).

We next examine the ACE2 expression in different types of cardiomyopathy. The most common types of cardiomyopathies includes dilated cardiomyopathy (DCM), hypertrophic cardiomyopathy (HCM), restrictive cardiomyopathy (RCM), arrhythmogenic right ventricular cardiomyopathy (ARVC) and left ventricular noncompaction (LVNC)^31–35^. These cardiomyopathy types have different causes and show distinctive morphology and physiology characteristics. Although previous studies have demonstrated ACE2 up-regulation in DCM and HCM^36,37^, how ACE2 is regulated in other types is unknown. Within the 7 datasets, we were able to identify all the common cardiomyopathy types. Our analysis revealed significantly increased ACE2 expression in most of the cardiomyopathy types (Fig. 3), including DCM, HCM, RCM, and LVNC. Although the result of ARVC is not statistically significant, the data show a clear trend of ACE2 upregulation (Fig. 3D).

**Figure 3:**
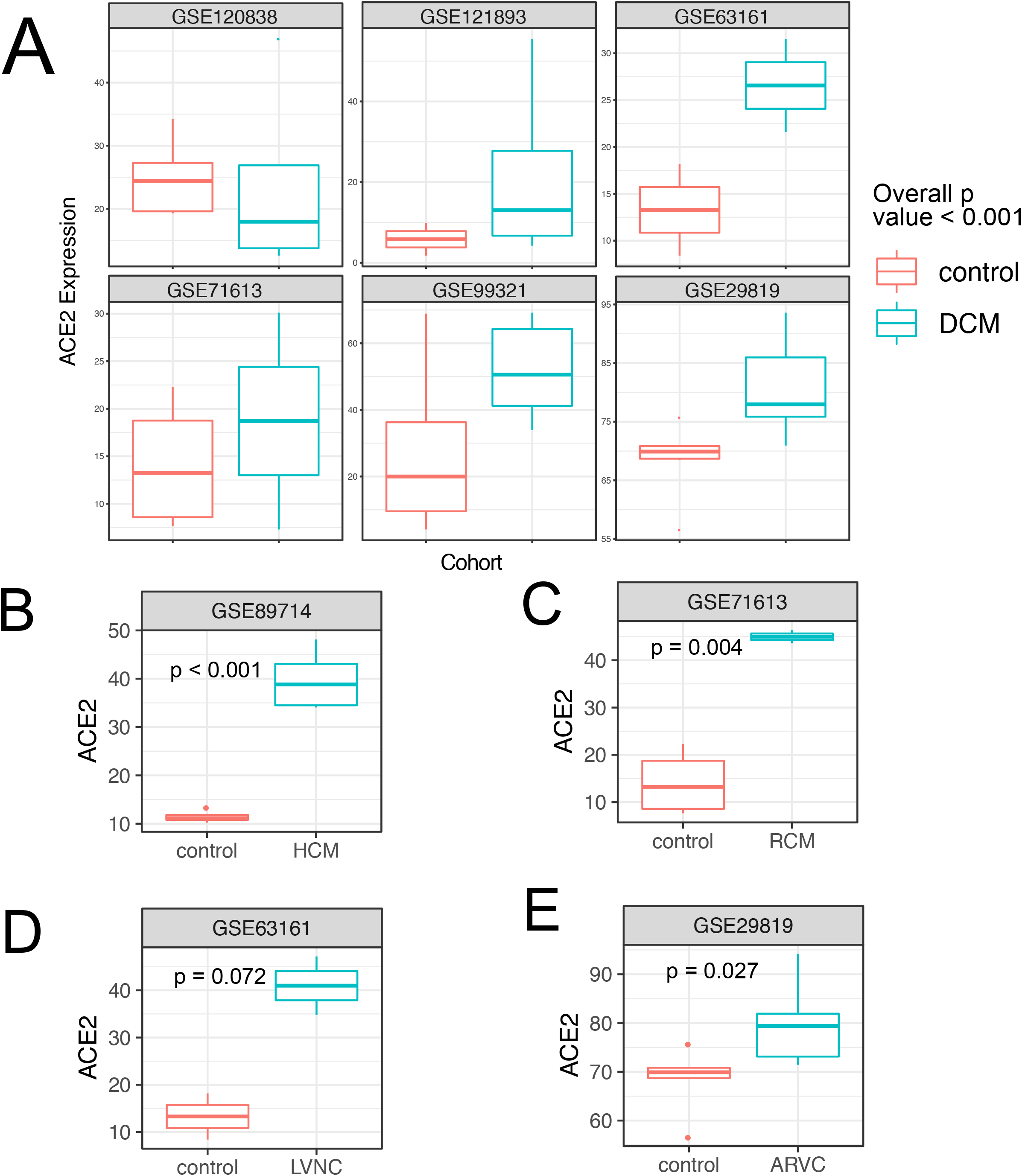
Meta-analysis confirms ACE2 upregulation in all major types of cardiomyopathy. (A) The ACE2 expression in normal hearts and hearts with DCM. Data are from five RNA-seq datasets and one microarray dataset. The overall p-value is calculated using a mixed model, with dataset as the random effect and DCM as the fixed effect. (B-E) The ACE2 expression in normal hearts and hearts with HCM, RCM, LVNC, and ARVC. p values in B-E are calculated using t-tests.

### COVID-19 patients with pre-existing cardiomyopathy show an increased mortality rate

While COVID19 patients with cardiovascular conditions show a higher mortality rate, it is not clear how cardiomyopathy, in particular, affects the survival of the patients. Because the ACE2 expression is significantly elevated in the heart of cardiomyopathy patients, we hypothesize that pre-existing cardiomyopathy leads to increased mortality in patients with COVID19.

We identified 3936 COVID19 patients from the electronic health records (EHR) of the University of California San Francisco (UCSF) hospital. We divided the patients into three groups, including patients with pre-existing cardiomyopathy (N = 43), patients with other pre-existing cardiovascular diseases (N = 624), and patients without cardiovascular diseases (N = 3269) (Table 2). The most common non-cardiomyopathy cardiovascular diseases include hypertension (N = 424), atherosclerotic heart diseases (N = 120) and Cardiac arrhythmia (N = 105).

**Table 2:** Demographic and clinical information of COVID-19 patients.

|  | Cardiomyopathy<br>(N=43) | Other cardiovascular<br>diseases<br>(N=624) | No cardiovascular<br>diseases<br>(N=3269) | Overall<br>(N=3936) |
| --- | --- | --- | --- | --- |
| **Demographics** |  |  |  |  |
| Age |  |  |  |  |
| Mean (SD) | 58.6 (17.9) | 55.9 (21.0) | 35.4 (22.0)*** | 38.9 (23.1) |
| Median [Min, Max] | 60.0 [18.0, 89.0] | 60.0 [0, 100] | 33.0 [0, 98.0] | 37.0 [0, 100] |
| Male | 33 (76.7%) | 296 (47.4%)* | 1481 (45.3%)*** | 1810 (46.0%) |
| Race |  |  |  |  |
| Asian | 12 (27.9%) | 103 (16.5%) | 489 (15.0%) | 604 (15.3%) |
| African American | 5 (11.6%) | 61 (9.8%) | 203 (6.2%) | 269 (6.8%) |
| MultiRace | 1 (2.3%) | 13 (2.1%) | 59 (1.8%) | 73 (1.9%) |
| Pacific Islander | 3 (7.0%) | 13 (2.1%) | 56 (1.7%) | 72 (1.8%) |
| Unknown | 9 (20.9%) | 142 (22.8%) | 1607 (49.2%) | 1758 (44.7%) |
| White | 13 (30.2%) | 289 (46.3%) | 849 (26.0%) | 1151 (29.2%) |
| Native American | 0 (0%) | 3 (0.5%) | 6 (0.2%) | 9 (0.2%) |
| **Pre-existing conditions** |  |  |  |  |
| Hyperlipidemia | 27 (62.8%) | 297 (47.6%) | 72 (2.2%)*** | 396 (10.1%) |
| Diabetes | 27 (62.8%) | 281 (45.0%) | 136 (4.2%)*** | 444 (11.3%) |
| Cancer | 10 (23.3%) | 145 (23.2%) | 67 (2.0%)*** | 222 (5.6%) |
| **Severe COVID19 presentations** |  |  |  |  |
| Ventilator use | 24 (55.8%) | 148 (23.7%)*** | 88 (2.7%)*** | 260 (6.6%) |
| Respiratory failure | 12 (27.9%) | 96 (15.4%) | 34 (1.0%)*** | 142 (3.6%) |
| Chest pain | 27 (62.8%) | 236 (37.8%)* | 140 (4.3%)*** | 403 (10.2%) |
| Mortality | 3 (7.0%) | 8 (1.3%) | 1 (0.0%)*** | 12 (0.3%) |
| Stars indicate the significant differences when compared the group to the cardiomyopathy group. * P< 0.05, ** P<0.01, *** P<0.001 |  |  |  |  |

We first compared the cardiomyopathy patients to patients without cardiovascular diseases. Patients with cardiomyopathy have a larger proportion of males and older ages. They also have a higher percentage of patients with preexisting conditions such as cancer, diabetes, and hyperlipidemia. A higher percentage of cardiomyopathy patients have severe COVID-19 disease presentations, including ventilator use, respiratory failure, chest pain, and death (Table 2). We then performed survival analysis to test the effect of cardiomyopathy while controlling for differences in age, gender, and pre-existing conditions using a multivariable Cox proportional-hazards model. We confirmed that cardiomyopathy is significantly associated with the risk of death (p = 0.004) (Figure 4A).

**Figure 4:**
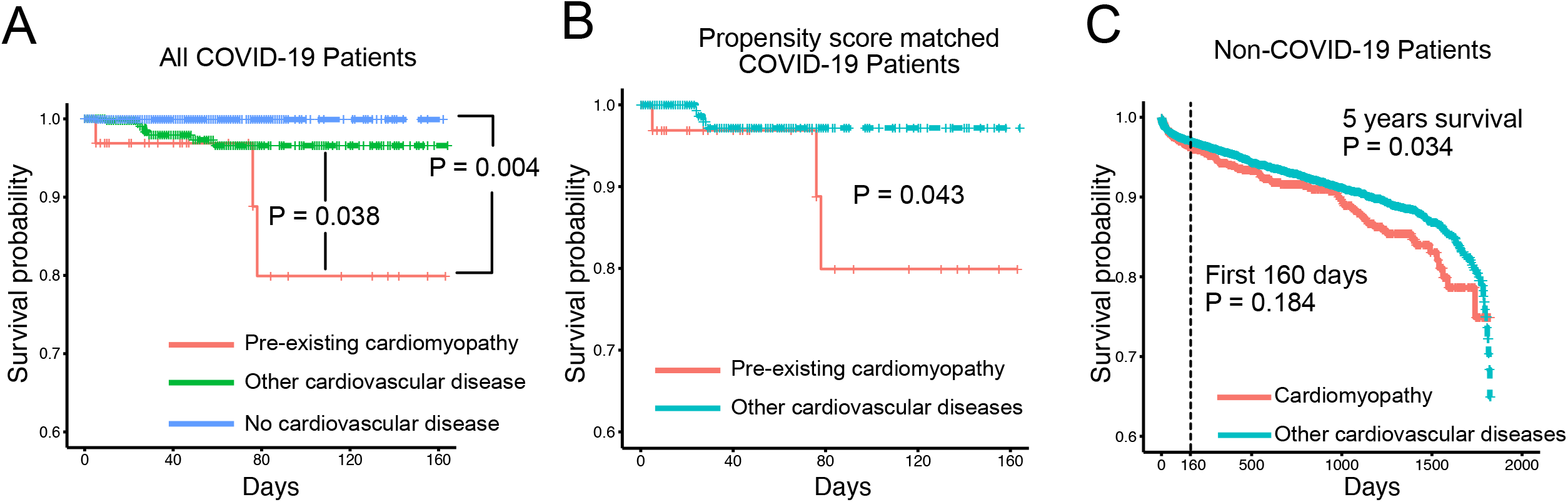
Patients with pre-existing cardiomyopathy show an increased mortality rate. (A) Kaplan Meier curve of COVID19 patients with pre-existing cardiomyopathy (N = 43), patients with other pre-existing cardiovascular diseases (N = 624), and patients without cardiovascular diseases (N = 3269). P values are from Cox proportional-hazards models, controlling for differences in the demographics and non-cardiovascular conditions between the groups. (B) Kaplan Meier curve of COVID19 patients with pre-existing cardiomyopathy (N = 43) and a cohort of propensity score-matched patients with other pre-existing cardiovascular diseases (N= 344). The P value is from a log-rank test. (C) Kaplan Meier curve of COVID-19 negative patients with pre-existing cardiomyopathy (N = 2250) and a cohort of propensity score-matched patients with other pre-existing cardiovascular diseases (N =18000). P values are from a log-rank test.

We next compared the cardiomyopathy patients to patients with other cardiovascular diseases. Cardiomyopathy patients have a higher proportion of males compared to patients with other cardiovascular diseases. Age, race, and pre-existing conditions are comparable between the two groups. Again, we observe that a higher percentage of cardiomyopathy patients have severe COVID-19 presentations, including ventilator use, chest pain, and death (Table 2). Multivariable Cox proportional-hazards regression confirms that cardiomyopathy is significantly associated with the risk of death (p = 0.038, 438 % increase in observed death rate) (Figure 4A). We further confirmed the increased mortality by comparing cardiomyopathy patients with a propensity score-matched cohort of patients with other cardiovascular diseases (Figure 4B and Table S1).

We then examined the survival of cardiomyopathy patients who are COVID-19 negative. We compared the survival of COVID-19 negative cardiomyopathy patients (N = 2250) with a propensity score-matched cohort of patients with other cardiovascular diseases (N = 18000). The two cohorts are comparable in demographics and non-cardiovascular diseases (Table S2). The 5-year mortality rate is only slightly higher in the cardiomyopathy patients (p = 0.034, 22% increase in observed death rate) (Fig. 4C). When we consider the patient’s survival at 160 days, a time frame comparable to the COVID-19 positive dataset, there is no significant difference between the survival of the two groups (Fig. 4C).

Taken together, the results show that cardiomyopathy itself does not pose large additional risk of mortality among patients with cardiovascular diseases. Rather, the interaction between SARS-CoV-2 infection and pre-existing cardiomyopathy leads to increased mortality in patients. Our transcriptomics analysis suggests that the up-regulated ACE2 expression may contribute to disease severity of COVID-19 in patients with pre-existing cardiomyopathy. However, further mechanistic studies are needed to establish the causal relationship between ACE2 up-regulation and the mortality in COVID-19 patients.

## Discussion

The disease severity of COVID-19 patients varies from asymptomatic to life-threatening. While we do not fully understand the reason behind such variation, it is clear that the disease severity is determined by multiple factors, including age, gender, the status of the immune system, and the pre-existing conditions^7,38–40^. ACE2 expression is a key determinant of the disease severity, as shown by multiple studies in humans and in animal models^3,5,9^. Therefore, it is critical to identify conditions that modulate ACE2 expression, as the information will help us reveal and explain factors associated with increased risk of severe illness from COVID-19.

We leveraged the massive amount of publicly available RNA-seq data to identify the ACE2 modulating conditions. While many tools exist for analyzing bulk GEO data, they are not optimized for this purpose. First, some tools require researchers to search for datasets using keywords, such as the name of a drug or a disease. These tools do not address our needs, as we are looking for any conditions that modulate ACE2 expression. Second, some tools require manual annotation of experimental groups in the studies, which are not scalable and often only cover a small subset of currently available datasets. Finally, the existing tools focus on differential expression analysis of two groups and are unable to address more complex experimental designs.

We address the problems using a variance analysis approach. Instead of comparing two experimental groups, we quantify the variance of the gene expression across all samples in a study. The rationale is that datasets with large ACE2 variances are likely to contain conditions that modulate the ACE2 expression. We improved our approach by using two modifications. First, we numerically embedded the metadata and calculated the regression coefficient R^2^ between ACE2 and the embedding. This allows us to prioritize datasets in which the ACE2 variation is associated with metadata. Second, we controlled for the overall heterogeneity of samples in the study, this allows us to prioritize datasets in which ACE2 are specifically modulated rather than as a result of cell-type differences.

Our study identifies multiple diseases, conditions, and genetic perturbations that modulate ACE2 expression. When interpreting these findings, readers should take into account several limitations of our study. First, many of the ACE2 modulating conditions are discovered based on data from one study with small sample sizes. These results should be viewed as data-driven hypotheses rather than definitive proofs. Additional data is required to confirm the findings. Second, our analysis does not establish a direct link between COVID-19 risk and the identified ACE2 modulators. Clinical studies are required to test if these ACE2 modulating conditions alter the risk of COVID-19 infection and pathogenesis. Finally, the datasets profile the ACE2 expression in bulk tissues. Therefore, it is not clear if the variation of ACE2 expression is due to a change in cellular composition or a change in transcriptional regulation. Single-cell analysis is required to identify the cause of ACE2 expression change.

Heart diseases are associated with severe COVID-19 illness through two mechanisms. Pre-existing heart conditions are comorbidities of COVID19^28,40^. On the other hand, SARS-COV-2 infection can induce acute myocardial injury^30^. Cardiomyopathy is one of the most common heart diseases. Compared to other age-related heart conditions, cardiomyopathy can affect individuals at any age. However, clinical studies were lacking to specifically characterize the disease severity of COVID-19 in patients with cardiomyopathy. Our transcriptional analysis highlights the significantly increased ACE2 expression in the hearts of cardiomyopathy patients, suggesting that cardiomyopathy patients are at an increased risk of heart damage caused by COVID-19.

Consistent with the RNA-seq finding, we found that COVID19 patients with pre-existing cardiomyopathy show increased mortality risk than other populations, even compared with age-matched populations with other cardiovascular conditions. While mechanistic research is needed to establish the causal relationship between ACE2 up-regulation and the increased mortality, our result identifies the cardiomyopathy patients as a high-risk group that needs extra protection and care.

This project demonstrated public RNA-sequencing data as a valuable resource for biomedical knowledge discovery. The existing RNA-seq data can be quickly repurposed to address pressing problems, such as identifying the risk factor of COVID19. While this study focused on analyzing the expression of ACE2, researchers can identify modulators for any genes or gene signatures of interest using the GENEVA web portal at genevatool.org.

## Methods

### Data preparation

We downloaded the uniformly processed RNA-seq data from ARCHS4 website (https://amp.pharm.mssm.edu/archs4/download.html) on August 03, 2020. The downloaded data include gene-level count data of 286650 samples from 9124 datasets and sample-level metadata. We transformed the gene count data into percentile rank data, which reduces the influences of library size, batch effects, and extreme values^41,42^. We downloaded study-level metadata using the entrez_search and entrez_summary function from the rentrez library^43^.

### Correlation analysis

We first calculated the Pearson correlation between ACE2 and other genes using data from all 286,650 samples. To calculate intra-study correlation, we selected a subset of studies in which the variance of ACE2 is greater than 10. We then calculated the Pearson correlation within each study. Transcription factors are identified by selecting genes with the Gene Ontology term “DNA-binding transcription factor activity (GO:0003700)”.

To identify pathways associated with ACE2, we used the pairwise correlations with ACE2 as a signature. We used the signature to query the Gene Ontology Biological Process database^44^. The fgsea function from the fgsea library was used to calculate the enrichment score^45^.

### Metadata embedding

We first concatenated the metadata of each sample into a single string, including the title, tissue type, and other characteristics (e.g. demographics, time points, treatment, genetic information, and disease status). We then calculated the pairwise Levenshtein distance between the strings that belong to the same study (GEO series). We applied multidimensional scaling to the pairwise Levenshtein distance and embedded the strings into 2-dimensional space for visualization and downstream analysis.

### GENEVA analysis

For a given gene in a given dataset, we first calculated the variance of the gene (VARg). We measure the overall heterogeneity of the samples by calculating the average variance of all genes (VARm). We run a regression using the expression of the gene as the dependent variable and the embedded metadata as independent variables (expression ∼ first embed dimension + second embed dimension). The regression coefficient (R^2^) represents the association between the expression of the gene and the embedded metadata. The product between VARg and R^2^ represents the variance of the gene explained by the embedded metadata. The GENEVA score is defined as VARg × R^2^ / VARm.

To test the significance of the GENEVA scores, we shuffled the samples across all datasets. We then calculated the GENEVA scores of all shuffled datasets to create a null distribution. Given a GENEVA score G, its p-value is defined as the probability that the null distribution is greater than G: p-value = Prob(null > G). We adjust the p values for multiple testing using the false discovery rate method.

### Meta-analysis

We searched the gene expression omnibus using the keyword “cardiomyopathy”. We then filter the results to only include studies that 1) profiled the transcriptome of heart tissues from humans and 2) compared cardiomyopathy samples with healthy samples. We identified 7 studies. We used a mixed effect model to test the effect of cardiomyopathy on ACE2 expression: ACE2 expression ∼ study (random effect) + cardiomyopathy status (fixed effect).

To examine the ACE2 expression in different types of cardiomyopathy, we separated the cardiomyopathy samples based on their subtype. We matched the cardiomyopathy samples with healthy controls within the same study. We used unpaired T-tests to test the effect of each cardiomyopathy type on ACE2 expression. Since data from multiple studies are available for dilated cardiomyopathy (DCM), we used a mixed effect model to test the effect of DCM on ACE2 expression: ACE2 expression ∼ study (random effect) + cardiomyopathy status (fixed effect).

### Analysis of electronic health records

The UCSF COVID-19 Data Mart records the clinical information of COVID19 patients and selected control patients using the Observational Medical Outcomes Partnership (OMOP) data format. We identified COVID19 patients from the clinical data using the ICD10 code U07. 1. We identified cardiomyopathy patients using the ICD10 codes I42 and I43, entered before their first COVID19 diagnosis. We identified patients with other cardiovascular diseases using the following ICD10 codes I00 - I99. We compared the cardiomyopathy patient with patients with other cardiovascular diseases and patients without cardiovascular diseases. We tested if the demographic and clinical variables are significantly different between the groups using single variable logistic regressions (cardiomyopathy ∼ clinical variable). We used the coxph function in the survival R package to perform the survival analysis, controlling for the variables that are significantly different in the logistic regressions. Patient survival time is defined as the time between their first COVID19 diagnosis and their death date. Live patients are censored on the last day of their encounter. In the COVID-19 negative cohort, cardiomyopathy patients are defined as patients whose first cardiovascular-related diagnosis is cardiomyopathy (ICD10 code I42 or I43). Patients with other cardiovascular diseases are defined as patients who have cardiovascular diseases (ICD10 code I00 - I99), but do not have cardiomyopathy. Patient survival time is defined as the time between their first cardiovascular disease and their death. Live patients are censored on the last day of their encounter. To perform propensity score matching, we first calculated the propensity score using logistic regression (cardiomyopathy ∼ age + race + gender + non-cardiovascular pre-existing conditions). We subsampled the non-cardiomyopathy cohort so that the distribution of its propensity score matches with the cardiomyopathy cohort.

### GENEVA web portal

GENEVA webportalGENEVA web tool is an open-source application available under GNU General Public License at http://genevatool.org. It is implemented in python web framework Django. The source code for the tool is available in the public Git repository at https://github.com/NavchetanKaur/geneva-webtool. The tool offers an intuitive interface and user guide on the home page. Users can select either of the two options from “Gene Query” and “Gene Signature Query” and query their gene of interest or set of upregulated and downregulated genes of interest. The results are displayed in tabular form with calculated GENEVA scores. GSE descriptions are further represented in plots and tables.

## Data Availability

The RNA-seq data is publicly available at Gene Expression Omnibus and can be explored using the GENEVA tool: genevatool.org

http://genevatool.org

## Acknowledgment

We thank all researchers who have contributed RNA-seq datasets to the Gene Expression Omnibus. We thank Dr. Alexander Lachmann, Dr. Avi Ma’ayan, and other co-authors for creating ARCHS4, which enabled our study. We thank Dr. Douglas Arneson and Sanchita Bhattacharya for helpful discussion. We thank the UCSF COVID-19 Data Mart for providing the electronic health record data of COVID-19 patients. This work was partially supported by the National Institute of Allergy and Infectious Diseases (Bioinformatics Support Contract HHSN272201200028C). The content is solely the responsibility of the authors and does not necessarily represent the official views of the National Institutes of Health. The analysis of the electronic health record data is conducted with approval from the UCSF institutional review board (IRB #: 20-31107).

## Conflicts of Interest

Atul Butte is a co-founder and consultant to Personalis and NuMedii; consultant to Samsung, Mango Tree Corporation, and in the recent past, 10x Genomics, Helix, Pathway Genomics, and Verinata (Illumina); has served on paid advisory panels or boards for Geisinger Health, Regenstrief Institute, Gerson Lehman Group, AlphaSights, Covance, Novartis, Genentech, Merck, and Roche; is a shareholder in Personalis and NuMedii; is a minor shareholder in Apple, Facebook, Alphabet (Google), Microsoft, Amazon, Snap, 10x Genomics, Illumina, CVS, Nuna Health, Assay Depot, Vet24seven, Regeneron, Sanofi, Royalty Pharma, AstraZeneca, Moderna, Biogen, Paraxel, and Sutro, and several other non-health related companies and mutual funds; and has received honoraria and travel reimbursement for invited talks from Johnson and Johnson, Roche, Genentech, Pfizer, Merck, Lilly, Takeda, Varian, Mars, Siemens, Optum, Abbott, Celgene, AstraZeneca, AbbVie, Westat, and many academic institutions, medical or disease specific foundations and associations, and health systems. Atul Butte receives royalty payments through Stanford University, for several patents and other disclosures licensed to NuMedii and Personalis. Atul Butte’s research has been funded by NIH, Northrup Grumman (as the prime on an NIH contract), Genentech, Johnson and Johnson, FDA, Robert Wood Johnson Foundation, Leon Lowenstein Foundation, Intervalien Foundation, Priscilla Chan and Mark Zuckerberg, the Barbara and Gerson Bakar Foundation, and in the recent past, the March of Dimes, Juvenile Diabetes Research Foundation, California Governor’s Office of Planning and Research, California Institute for Regenerative Medicine, L’Oreal, and Progenity.

**Supplemental Figure 1:**
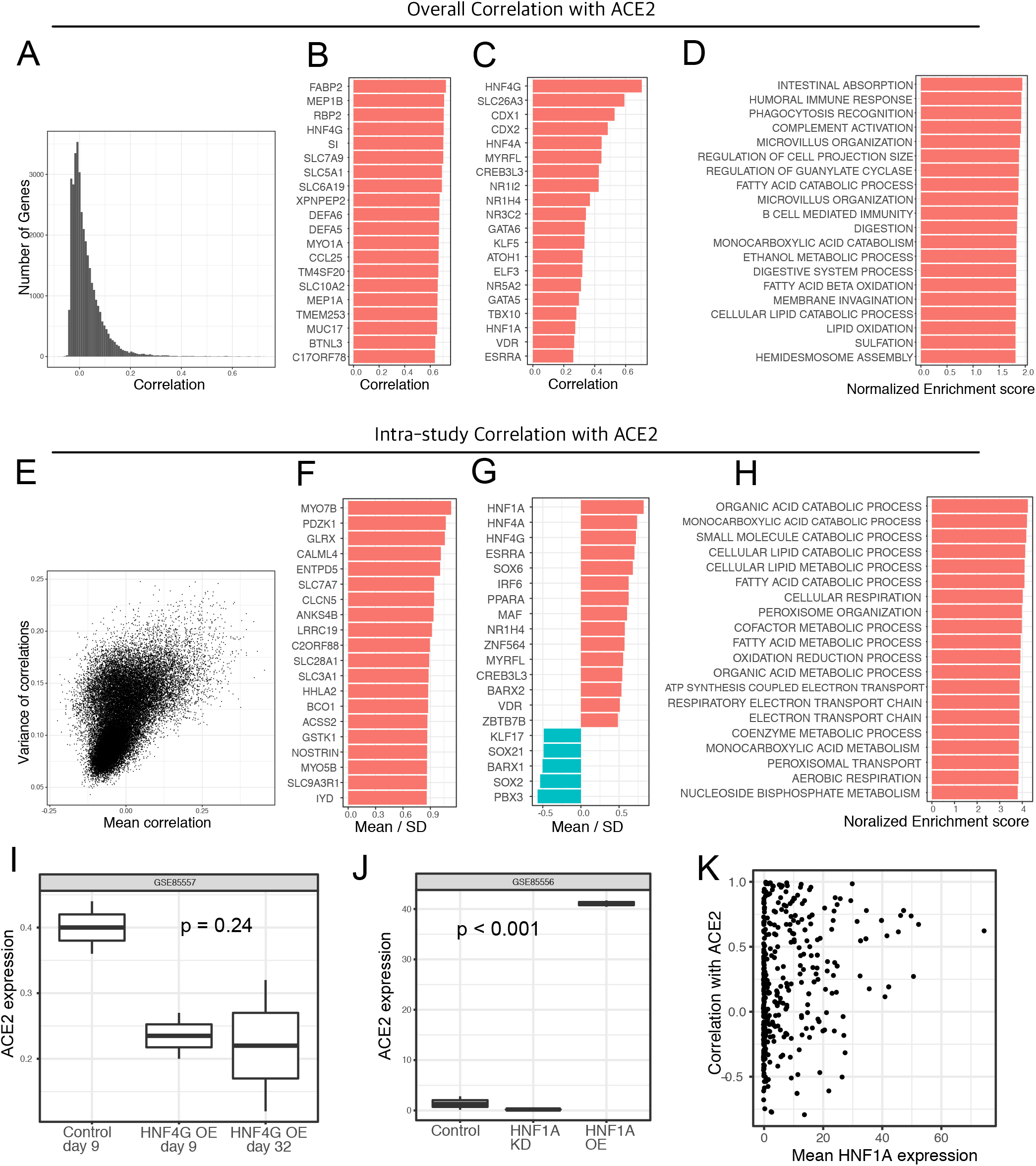
RNA-seq analysis reveals complex transcriptional networks of ACE2. We calculated the pairwise Pearson correlations between ACE2 and other genes using all data from 286650 RNA-seq samples (A-D). (A) Histogram of the pairwise correlations. (B) Top 30 genes with the highest correlation with ACE2. (C) Top 30 transcription factors with the highest correlation with ACE2. (D) Top 30 pathways correlated with ACE2 expression. We calculated the pairwise Pearson correlations between ACE2 and other genes using samples in individual datasets (E-H). (E) Scatter plot shows the mean and variance of the correlation between ACE2 and other genes across different datasets. (F) Top 30 genes with the highest standardized correlation with ACE2. The standardized correlation is defined as the average correlation divided by its standard deviation. (G) Top 30 transcription factors with the highest standardized correlation with ACE2. (H) Top 30 pathways with the highest standardized correlation with ACE2. (I) Box plot showing the expression of ACE2 in LNCaP/AR cells with or without HNF4G over expression. (I) Box plot showing the expression of ACE2 in LNCaP/AR cells with or without HNF4A over expression or knock down. (K) scatter plot shows the relationship between HNF4G expression level and its correlation with ACE2.

**Table S1:** Demographic and clinical information of COVID-19 patients with cardiomyopathy and a propensity score matched cohort of COVID-19 patients with other cardiovascular diseases.

|  | pre-existing<br>cardiomyopathy<br>(N=43) | Other cardiovascular<br>diseases<br>(N=344) | Overall<br>(N=387) |
| --- | --- | --- | --- |
| **age** |  |  |  |
| Mean (SD) | 58.6 (17.9) | 57.9 (19.7) | 58.0 (19.5) |
| Median [Min, Max] | 60.0 [18.0, 89.0] | 60.5 [0, 100] | 60.0 [0, 100] |
| **gender** |  |  |  |
| Female | 10 (23.3%) | 95 (27.6%) | 105 (27.1%) |
| Male | 33 (76.7%) | 249 (72.4%) | 282 (72.9%) |
| **race** |  |  |  |
| Asian | 12 (27.9%) | 77 (22.4%) | 89 (23.0%) |
| Black or African American | 5 (11.6%) | 33 (9.6%) | 38 (9.8%) |
| MultiRace | 1 (2.3%) | 9 (2.6%) | 10 (2.6%) |
| Native Hawaiian or Other Pacific Islander | 3 (7.0%) | 13 (3.8%) | 16 (4.1%) |
| Unknown | 9 (20.9%) | 74 (21.5%) | 83 (21.4%) |
| White | 13 (30.2%) | 138 (40.1%) | 151 (39.0%) |
| **hyperlipidemia** |  |  |  |
| Yes | 27 (62.8%) | 195 (56.7%) | 222 (57.4%) |
| No | 16 (37.2%) | 149 (43.3%) | 165 (42.6%) |
| **diabetes** |  |  |  |
| Yes | 27 (62.8%) | 184 (53.5%) | 211 (54.5%) |
| No | 16 (37.2%) | 160 (46.5%) | 176 (45.5%) |
| **cancer** |  |  |  |
| Yes | 10 (23.3%) | 81 (23.5%) | 91 (23.5%) |
| No | 33 (76.7%) | 263 (76.5%) | 296 (76.5%) |
| **ventilator use** |  |  |  |
| Yes | 24 (55.8%) | 90 (26.2%) | 114 (29.5%) |
| No | 19 (44.2%) | 254 (73.8%) | 273 (70.5%) |
| **respiratory failure** |  |  |  |
| Yes | 12 (27.9%) | 62 (18.0%) | 74 (19.1%) |
| No | 31 (72.1%) | 282 (82.0%) | 313 (80.9%) |
| **chest pain** |  |  |  |
| Yes | 27 (62.8%) | 125 (36.3%) | 152 (39.3%) |
| No | 16 (37.2%) | 219 (63.7%) | 235 (60.7%) |
| **death** |  |  |  |
| Yes | 3 (7.0%) | 4 (1.2%) | 7 (1.8%) |
| No | 40 (93.0%) | 340 (98.8%) | 380 (98.2%) |

**Table S2:** Demographic and clinical information of Non-COVID-19 patients with cardiomyopathy and a propensity score matched cohort of patients with other cardiovascular diseases.

|  | Cardiomyopathy<br>(N=2250) | Other cardiovascular diseases<br>(N=18000) | Overall<br>(N=20250) |
| --- | --- | --- | --- |
| **age** |  |  |  |
| Mean (SD) | 45.9 (23.3) | 46.4 (24.1) | 46.3 (24.0) |
| Median [Min, Max] | 50.0 [0, 90.0] | 51.0 [0, 91.0] | 51.0 [0, 91.0] |
| **gender** |  |  |  |
| FEMALE | 897 (39.9%) | 7238 (40.2%) | 8135 (40.2%) |
| MALE | 1352 (60.1%) | 10758 (59.8%) | 12110 (59.8%) |
| UNKNOWN | 1 (0.0%) | 4 (0.0%) | 5 (0.0%) |
| **race** |  |  |  |
| American Indian or Alaska Native | 9 (0.4%) | 73 (0.4%) | 82 (0.4%) |
| Asian | 200 (8.9%) | 1526 (8.5%) | 1726 (8.5%) |
| Black or African American | 190 (8.4%) | 1539 (8.6%) | 1729 (8.5%) |
| MultiRace | 122 (5.4%) | 1018 (5.7%) | 1140 (5.6%) |
| Native Hawaiian or Other Pacific Islander | 27 (1.2%) | 195 (1.1%) | 222 (1.1%) |
| Other Race | 382 (17.0%) | 3172 (17.6%) | 3554 (17.6%) |
| Unknown | 335 (14.9%) | 2449 (13.6%) | 2784 (13.7%) |
| White | 985 (43.8%) | 8028 (44.6%) | 9013 (44.5%) |
| **hyperlipidemia** |  |  |  |
| Yes | 368 (16.4%) | 2945 (16.4%) | 3313 (16.4%) |
| No | 1882 (83.6%) | 15055 (83.6%) | 16937 (83.6%) |
| **diabetes** |  |  |  |
| Yes | 370 (16.4%) | 2874 (16.0%) | 3244 (16.0%) |
| No | 1880 (83.6%) | 15126 (84.0%) | 17006 (84.0%) |
| **cancer** |  |  |  |
| Yes | 306 (13.6%) | 2430 (13.5%) | 2736 (13.5%) |
| No | 1944 (86.4%) | 15570 (86.5%) | 17514 (86.5%) |
| **death** |  |  |  |
| Yes | 123 (5.5%) | 807 (4.5%) | 930 (4.6%) |
| No | 2127 (94.5%) | 17193 (95.5%) | 19320 (95.4%) |

## Notes

### Author Declarations

The analysis of the electronic health record data is conducted with approval from the UCSF institutional review board (IRB #: 20-31107).

